# Incorporating human mobility data improves forecasts of Dengue fever in Thailand

**DOI:** 10.1101/2020.07.22.20157966

**Authors:** Mathew V Kiang, Mauricio Santillana, Jarvis T Chen, Jukka-Pekka Onnela, Nancy Krieger, Kenth Engø-Monsen, Nattwut Ekapirat, Darin Areechokchai, Preecha Prempree, Richard J. Maude, Caroline O Buckee

## Abstract

Over 390 million people worldwide are infected with dengue fever each year. In the absence of an effective vaccine for general use, national control programs must rely on hospital readiness and targeted vector control to prepare for epidemics, so accurate forecasting remains an important goal. Many dengue forecasting approaches have used environmental data linked to mosquito ecology to predict when epidemics will occur, but these have had mixed results. Conversely, human mobility, an important driver in the spatial spread of infection, is often ignored. Here we compare time-series forecasts of dengue fever in Thailand, integrating epidemiological data with mobility models generated from mobile phone data. We show that long-distance connectivity is correlated with dengue incidence at forecasting horizons of up to three months, and that incorporating mobility data improves traditional time-series forecasting approaches. Notably, no single model or class of model always outperformed others. We propose an adaptive, mosaic forecasting approach for early warning systems.

## Introduction

More than half the world’s population is at risk of infection from the dengue virus, which causes an estimated 390 million infections (Bhatt et al., 2013) and 25,000 deaths per year (CDC, 2018; Guzman and Harris, 2015; WHO, 2018a). The dengue pathogen is spread in urban and peri-urban areas by invasive mosquitoes belonging to the Aedes complex. As a result, dengue has emerged as a major threat in the context of a rapidly urbanizing, globally connected world (Guzman and Harris, 2015; Tatem et al., 2006; Wesolowski et al., 2015b). For example, despite the general decline in the incidence of other communicable diseases, the incidence of dengue fever has doubled every 10 years since 1990 (Stanaway et al., 2016). The rapid geographic expansion of the vector suggests there will be a continuing emergence of dengue globally (Guzman and Harris, 2015; Tatem et al., 2006; Wesolowski et al., 2015b). Currently, there is no drug treatment for dengue (Halstead, 2012; WHO, 2012) and only a partially effective vaccine, which cannot be used in seronegative individuals (WHO, 2018b). Therefore, despite the mixed results of vector control efforts (WHO, 2012), targeted and thorough vector control approaches, hospital readiness, and risk communication can improve public health preparedness for seasonal outbreaks. Fundamental to the success of these preparations is data on the burden of disease in different areas, and some sense of how an epidemic may progress in the near term and on local spatial scales relevant to national control programs.

Forecasting the epidemic trajectory of dengue on weekly or monthly timescales remains a relatively new science for infectious diseases (Baquero et al., 2018; Buczak et al., 2018; Choudhury et al., 2008; Eastin et al., 2014; Gharbi et al., 2011; Hii et al., 2012; Hu et al., 2010; Johansson et al., 2016; Lauer et al., 2018; Martinez et al., 2011; Promprou et al., 2006; Nicholas G. Reich et al., 2016; Yamana et al., 2016; Yang et al., 2017). Unlike weather and climate forecasting, where physical laws dictate the dynamics of the system, the social and biological dynamics that drive infectious disease outbreaks make forecasting dengue epidemics challenging. Recurring epidemics, as opposed to novel pathogens emerging for the first time, occur against a backdrop of shifting population immunity, which is difficult to quantify. Complicating surveillance, pathogens like dengue are primarily reported based on symptoms rather than laboratory confirmation. Like influenza and malaria, dengue causes non-specific symptoms, fever in particular, so reporting reliability and time lags impact data quality (Chretien et al., 2016; Olliaro et al., 2018; Scarpino et al., 2017). Despite these complexities, routine forecasting is an important priority for national dengue control programs (Nicholas G. Reich et al., 2016; WHO, 2012).

There has been a recent surge of interest and success in building forecasting models for seasonal epidemics of dengue fever (Choudhury et al., 2008; Eastin et al., 2014; Gharbi et al., 2011; Hii et al., 2012; Hu et al., 2010; Johansson et al., 2016; Lauer et al., 2018; Martinez et al., 2011; Promprou et al., 2006; Nicholas G. Reich et al., 2016; Yamana et al., 2016; Yang et al., 2017). A distinction can be made between mechanistic epidemiological models and statistical models. Mechanistic models in which the mode of transmission (in this case, mosquito-borne and strong temperature dependence) is built into the model and drives the predicted infection dynamics. In contrast, statistical models rely on the identification of past epidemiological activity patterns and historical correlations with external data streams, generated often by human behavior on Internet search engines or social media, to monitor disease activity and predict future outbreaks. Mechanistic models aim at providing biological insight and a basis for interpretation, but for socially and environmentally complex infections like dengue, these models are often challenging to parameterize. Dengue is particularly challenging as it is composed of multiple immunologically distinct strains and relies on the interaction of mosquito and human population dynamics and microclimate variability. Metapopulation models have been developed to incorporate the spatial dynamics of dengue outbreaks, modeling each area with a set of location-specific parameters and linking the areas through estimated migration of individuals. Metapopulation models play in an important role in our understanding of epidemic outbreaks across spatial regions (Arino and Driessche, 2003; Liu et al., 2018; Stolerman et al., 2015), synchronicity between regions (Lloyd and Jansen, 2004), oscillations of epidemics (Lourenço and Recker, 2013), and strategies to reduce transmission (Lee and Castillo-Chavez, 2015). Despite their importance in understanding dynamics, mechanistic models, and metapopulation models in particular, may lack sufficient data for appropriate parameterization, and are often not feasible in a forecasting context. As a result, statistical models have been more successful for outbreak preparedness for which the modeling goal is to provide quantitative, relatively short-term predictions with explicit uncertainty (11, 13–20, 22, 28–30).

Most statistical forecasting approaches for dengue have been based on autocorrelation in case data, often incorporating environmental information due to the importance of temperature and other factors to the availability of mosquitoes and variation of the incubation period of the virus in the vector. Many of these have focused on long-term predictions of dengue at the city level (Choudhury et al., 2008; Eastin et al., 2014; Luz et al., 2008; Stolerman et al., 2016), or larger regions within a specific country (Gharbi et al., 2011; Hu et al., 2010; Martinez et al., 2011; Promprou et al., 2006). Models often show mixed success with high prediction accuracy in the immediate forecasting horizons (e.g., 1-2 months) and rapid decay at longer time horizons (e.g. 3-6 months). It is unclear if weather or climate variables substantially improve forecasting; at least one study that systematically looked at different model parameters for autoregressive models, with and without a wide range of climate variables, across states in Mexico found no conclusive improvement (Johansson et al., 2016). More recently, ensemble models have become a powerful way to combine different approaches in order to leverage the strengths of each while minimizing the weaknesses (52). This approach has recently been applied to dengue (Yamana et al., 2016). Others have incorporated new sources of data from internet search terms to predict dengue nationally (Yang et al., 2017), employed novel statistical methods to predict dengue in San Juan, Puerto Rico (Ray et al., 2017), or combined common climate covariates with generalized additive models to predict annual incidence of dengue hemorrhagic fever (Lauer et al., 2018).

Although dengue outbreaks spread primarily via human travel, incorporating this aspect of the spatial connectivity between locations within forecasting frameworks has been challenging. Current forecasting models, both mechanistic and statistical, either ignore or make crude assumptions about how populations are connected by travel. Parameterizing human mobility is challenging due to a paucity of relevant data streams, particularly in low-income settings. We have previously used mobile phone records to quantify national movements and showed that they provide improved prediction for dengue outbreaks in Pakistan (Wesolowski et al., 2015b). Specifically, we used a gravity models to parametrize human mobility in a mechanistic framework because dengue was emerging into naïve populations, where statistical methods could not be used. Others have used daily commuting data to model mobility using a radiation model, which in turn is used to parameterize a mechanistic model (Zhu et al., 2016). Although considerable difficulty remains in accessing mobile phone records or other scalable data sources about mobility, it is clear that gravity models, radiation models, and other proxies for travel measures may perform poorly in many settings (Wesolowski et al., 2015a).

To date, almost all efforts to forecast dengue have either focused on optimizing a single modeling framework across regions, fitting parameters individually, or analyzed multiple models for a particular location. Few statistical models used for forecasting dengue incorporate spatial dependencies and none incorporate information about mobility patterns. Here, we contribute to the existing literature by using seven years of monthly dengue data (2010–2016) from Thailand, which has a developed dengue surveillance program, and mobility data from approximately 11 million mobile phone subscribers to show that long-distance travel is associated with correlated epidemiological cases. We compare model structures incorporating time-series approaches or spatial dependencies, and mobility data, finding that this improves model prediction, but no individual approach provides the best performing model in all locations over all time horizons.

We quantify the error for each province in Thailand, showing that provinces in the north of the country are more difficult to forecast with confidence than those in the south, regardless of model choice, and that different models’ performances may be linked to demographic and social factors such as population density and gross provincial product per capita. We propose that mosaic forecasting approaches, which dynamically adapt over time and space, and end up using the best model for that location and time period, are likely to be the most effective for use in early warning systems in national control programs.

## Results

### Greater than expected long-distance travel to and from Bangkok

To assess inter-province migration, we analyzed the call data records (CDR) of approximately 11 million mobile phone subscribers between August 1, 2017 and October 19, 2017. At the time of data collection, the mobile phone operator had about 26% of the market share and was the third largest provider in Thailand. Since travel patterns remained stable over our period of observation (coefficient of variation: 1.3%; SI Appendix, Fig. S1), we calculated average daily journeys between all pairs of provinces in both directions, and compared observed mobility in the CDR data to expected mobility based on gravity models (see Materials and Methods) assuming travel over our time period is consistent with travel for the rest of the year (SI Appendix, Fig. S2). We found that the routes of travel that deviate significantly from gravity model-based predictions in both directions are focused on Bangkok (Figure 1), with more travel than expected from long distances around the country such as Phuket and Bangkok itself (Figure 1, left), and less travel than expected within and around the city (Figure 1, right). These hot and cold spots, where higher or lower than expected travel was observed, were robust to the gravity model coefficients used (SI Appendix, Table S1).

**Figure 1.**
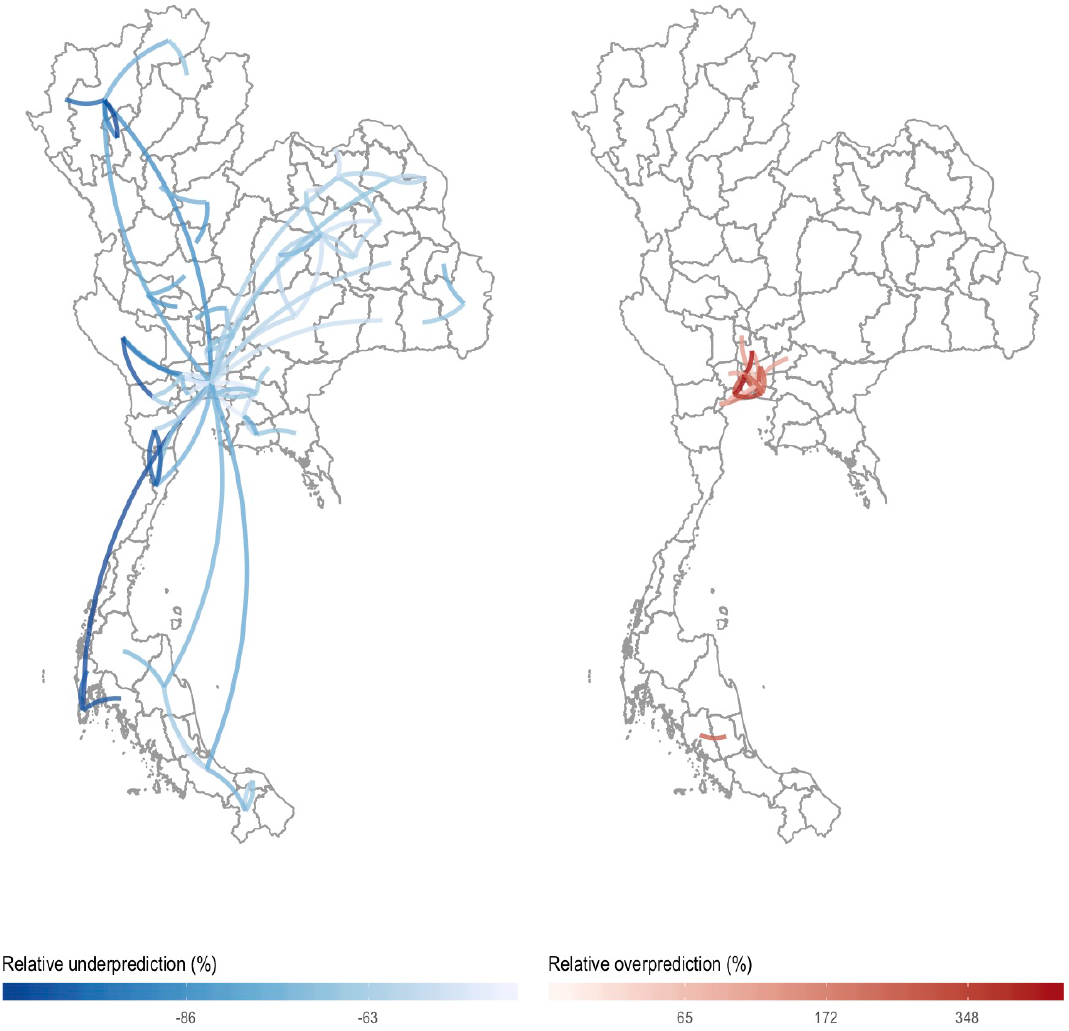
Under- and over-prediction of outlier travel. Relative under-prediction (left) and over-prediction (right) comparing observed mobility data (from CDRs) to estimated mobility data from the best fit gravity model. We defined relative prediction error as 100%*(PredictedTrips – ObservedTrips)/ObservedTrips. We highlight only observations with Cook’s distance greater than five times the average Cook’s distance. Note that Bangkok (center of the map) is central to much of the over- and under-prediction outliers with most over- prediction near Bangkok.

### Long-distance connectivity is associated with correlated dengue incidence

In Thailand, dengue follows a seasonal cycle across all 77 provinces (Figure 2), with variation in the timing of onset and epidemic peak in different locations over our period of observation (Limkittikul et al., 2014). We analyzed the correlation between clinical cases in each province with different time lags between them. Figure 3 shows the relationship between the correlation in dengue cases between pairs of provinces, stratified with respect to geographic distance and mobility measured using mobile phone data. Consistent with previous studies (Cummings et al., 2004; Panhuis et al., 2015; Salje et al., 2017, 2012), the epidemiological correlation between provinces is strongest when they are close to each other and declines with distance and over time (i.e. the three-month lagged correlation is weaker than the one-month lagged correlation). For provinces less than 1,000 km apart, human mobility estimated using mobile phone data does not appear to impact the correlation of clinical cases. For longer distances, however, more highly connected locations show higher correlation in clinical dengue cases than locations the same distance apart but with low observed connectivity (Figure 3). Note that some but not all of these long-distance connections are locations with international airports (SI Appendix, Fig. S3), and provinces connected by airports have higher correlation than those that are not connected by airports (SI Appendix, Fig. S4).

**Figure 2.**
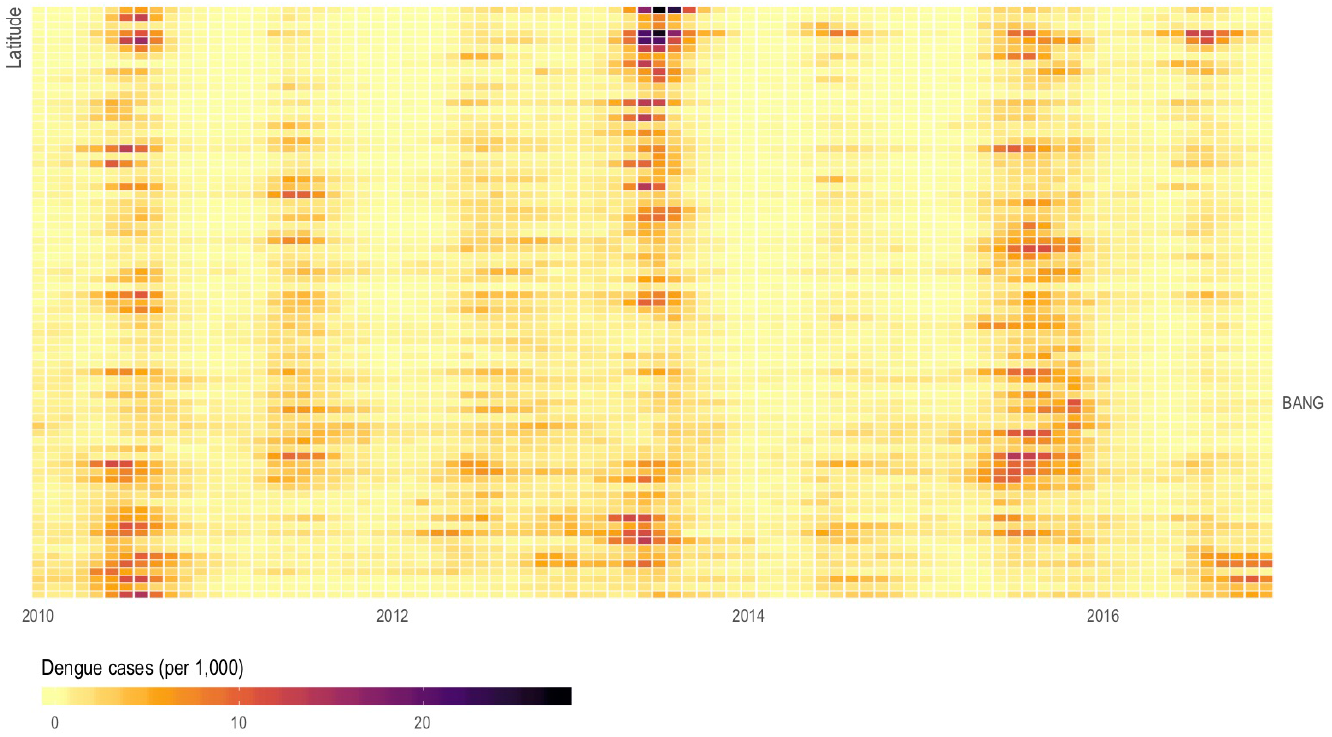
Monthly dengue incidence by province. Monthly crude incidence of dengue (per 1,000 person-years) by province (y-axis) ordered by centroid latitude (higher is more northern) over seven years of observation (x-axis). Dengue in Thailand follows a seasonal cycle with geographic variation in both the timing of onset and peak of the epidemic.

**Figure 3.**
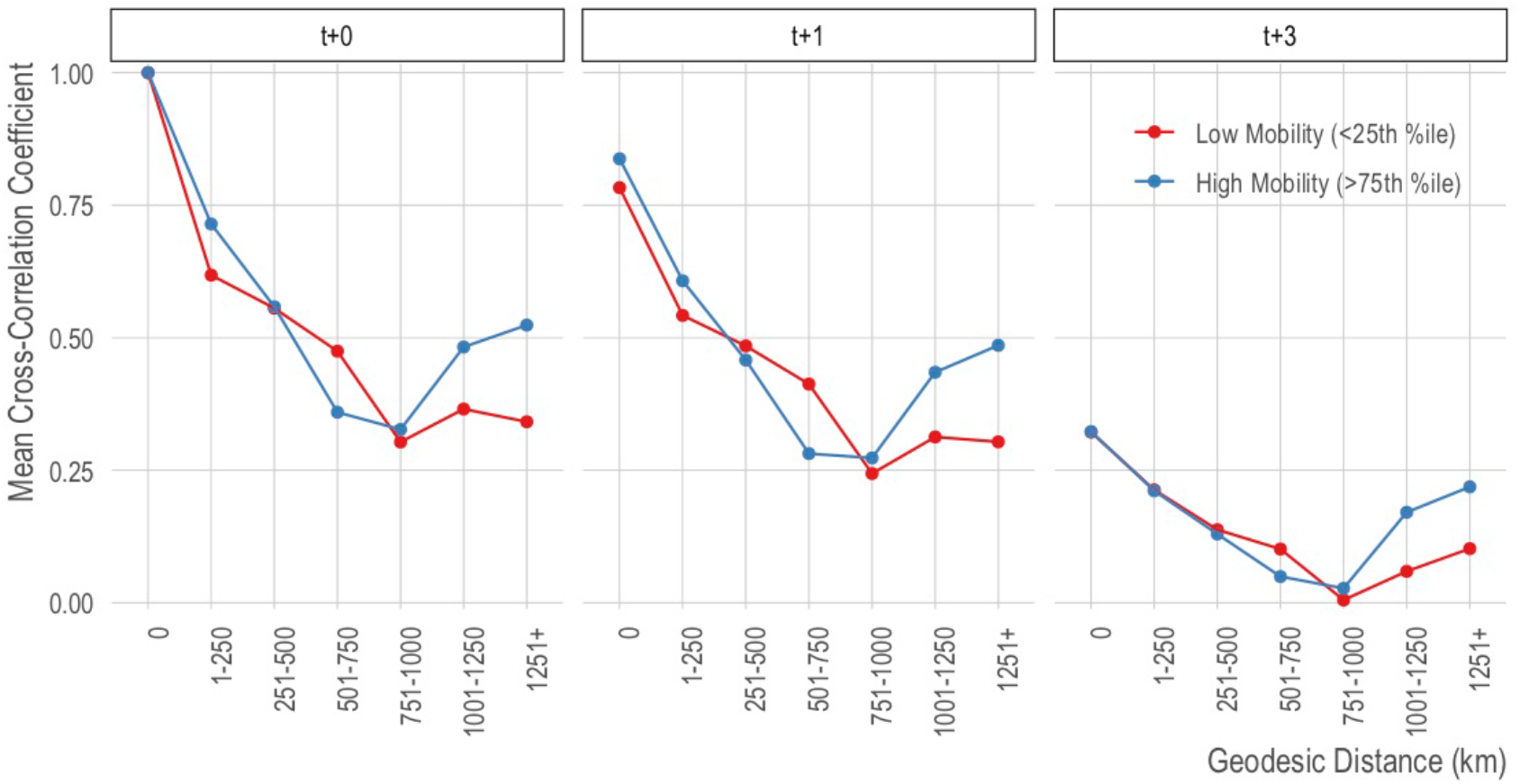
Correlation of province-level dengue by distance, at different time lags. We show the mean cross-correlation coefficient (y-axis) for pairs of provinces at binned distances (x-axis; 0 indicates correlation of an area with itself) for synchronous dengue (left panel) and lagged by 1 month (middle panel) and three months (right panel). The red line shows the bottom quartile of provinces in terms of incoming and outgoing travel and the blue line shows the top quartile.

### No one-size-fits-all: forecasting performance varies in space and time

We compared several forecasting approaches for the 77 Thai provinces to assess how model performance varied by region and over time, and to measure the impact of integrating the mobility data. Specifically, for each province, we fit four models: (1) local (non-spatially dependent) models commonly used for dengue; specifically, seasonal autoregressive integrated moving average models (Plain SARIMA) across a grid of parameters, (2) SARIMA models that use information from the top five most connected provinces (in terms of number of incoming trips) based on mobile phone data (CDR SARIMA), (3) SARIMA models that use information from the top five most connected provinces (in terms of predicted number of incoming trips) based on our gravity model estimates (Gravity SARIMA), and (4) a data-driven network approach, based on a regularized regression approach, that predicts dengue incidence in a given location potentially using dengue incidence from every other location as input (LASSO; see Materials and Methods and reference (49) for details).

Figure 4 illustrates the results of all models at all forecasting horizons for Bangkok (see SI Appendix, Text S1 for online-only results for all other provinces). At early forecasting horizons (i.e., one-month and up to three-months ahead time horizons), all models performed well, with the CDR SARIMA and Gravity SARIMA models outperforming the Plain SARIMA models by about 5–10% (Figure 5) as captured by the mean absolute error. After the 3-month ahead forecasting horizon, the Plain SARIMA model performance drops substantially faster than all other models. Importantly, the grouping of out-of-sample prediction errors, across forecasting horizons, tended to be closer in the LASSO models, indicating that across forecasting horizons, the network models lose predictive power more slowly than the SARIMA-based models. We present all plots for all provinces in an online repository (SI Appendix, Text S1).

**Figure 4.**
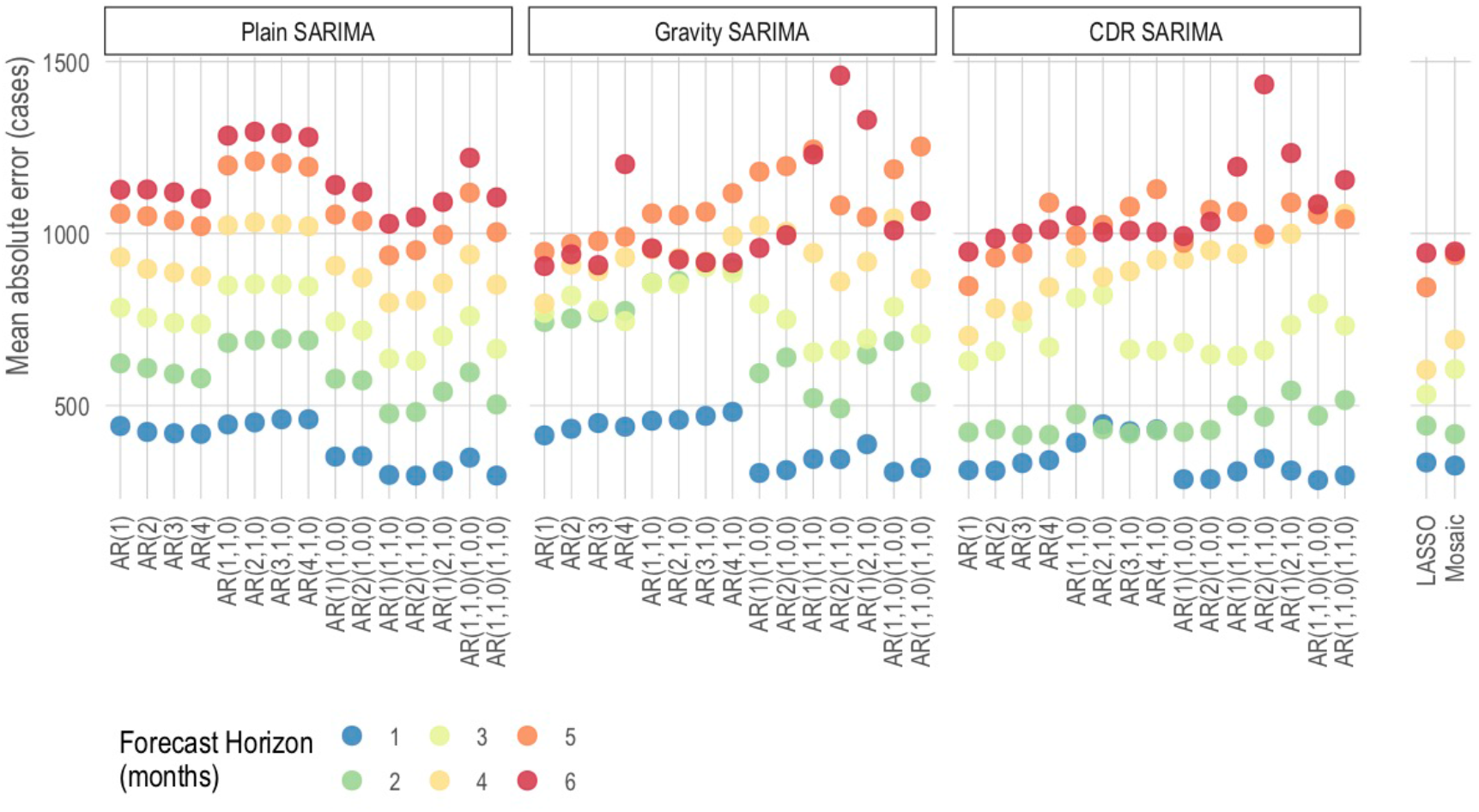
Mean absolute error (MAE) for all Bangkok models. The mean absolute error (y-axis) expressed as number of cases for each model (x-axis) and for each forecast. Models are grouped as SARIMA with no exogenous variables (Plain), SARIMAs with the top 5 most connected regions based on the predicted trips from a gravity model (Gravity SARIMA), and SARIMAs with the top 5 most connected regions based on CDR data (CDR SARIMA). The rightmost models show a data-driven network model, denoted as LASSO, since it is based on a least absolute shrinkage and selection operator prediction model, and mosaic model.

**Figure 5.**
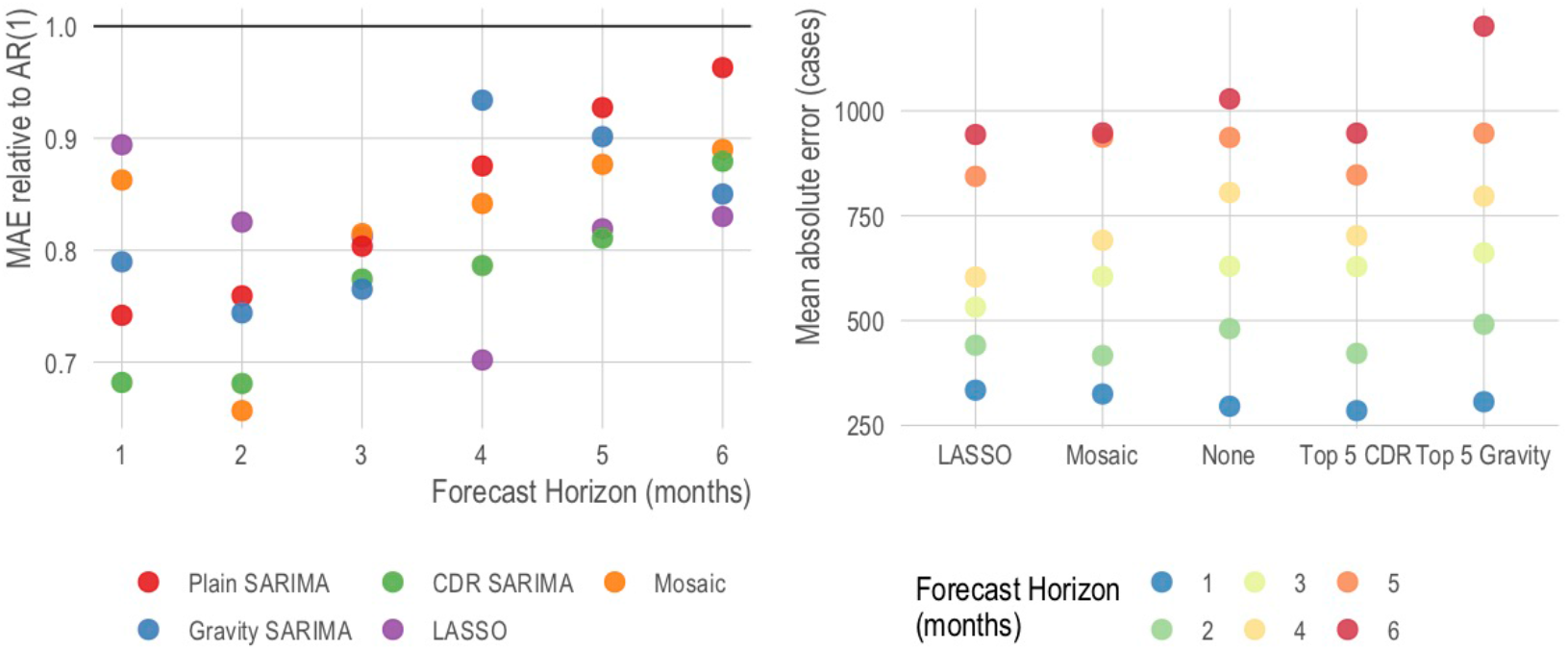
Comparing the best models for Bangkok, by model type. Focusing only on the best performing model for each model type and each time horizon, we show the relative mean absolute error (left panel) and the mean absolute error (right panel). On the left, the baseline of comparison is the traditional AR(1) model and the y-axis can be interpreted as the improvement over this baseline — i.e., a value of .9 indicates a 10% improvement. We show that both the Plain SARIMA (red) and CDR SARIMA (green) models perform better than the LASSO model at earlier forecasting horizons but perform worse at later horizons.

In general, no single model or class of model outperformed others across all provinces or all forecasting horizons (Figure 6; SI Appendix, Fig. S5). We found that across all model types, provinces in the south of the country had lower prediction errors compared to those in the north of the country (Figure 6). This difference in forecasting power was particularly pronounced on longer time horizons. For example, when comparing the out-of-sample prediction errors of the CDR SARIMA to the Plain SARIMA, the CDR SARIMAs were worse in 8 tasks for forecasting horizons of 1 to 3 months and better in only 3 tasks with no statistically significant difference in the remaining 220 prediction tasks. However, for forecasting horizons of 4-6 months, the CDR SARIMA outperformed the Plain SARIMA in 40 tasks and only underperformed in 8 with no statistically significant difference in the remaining 183 tasks (SI Appendix, Fig. S6).

**Figure 6.**
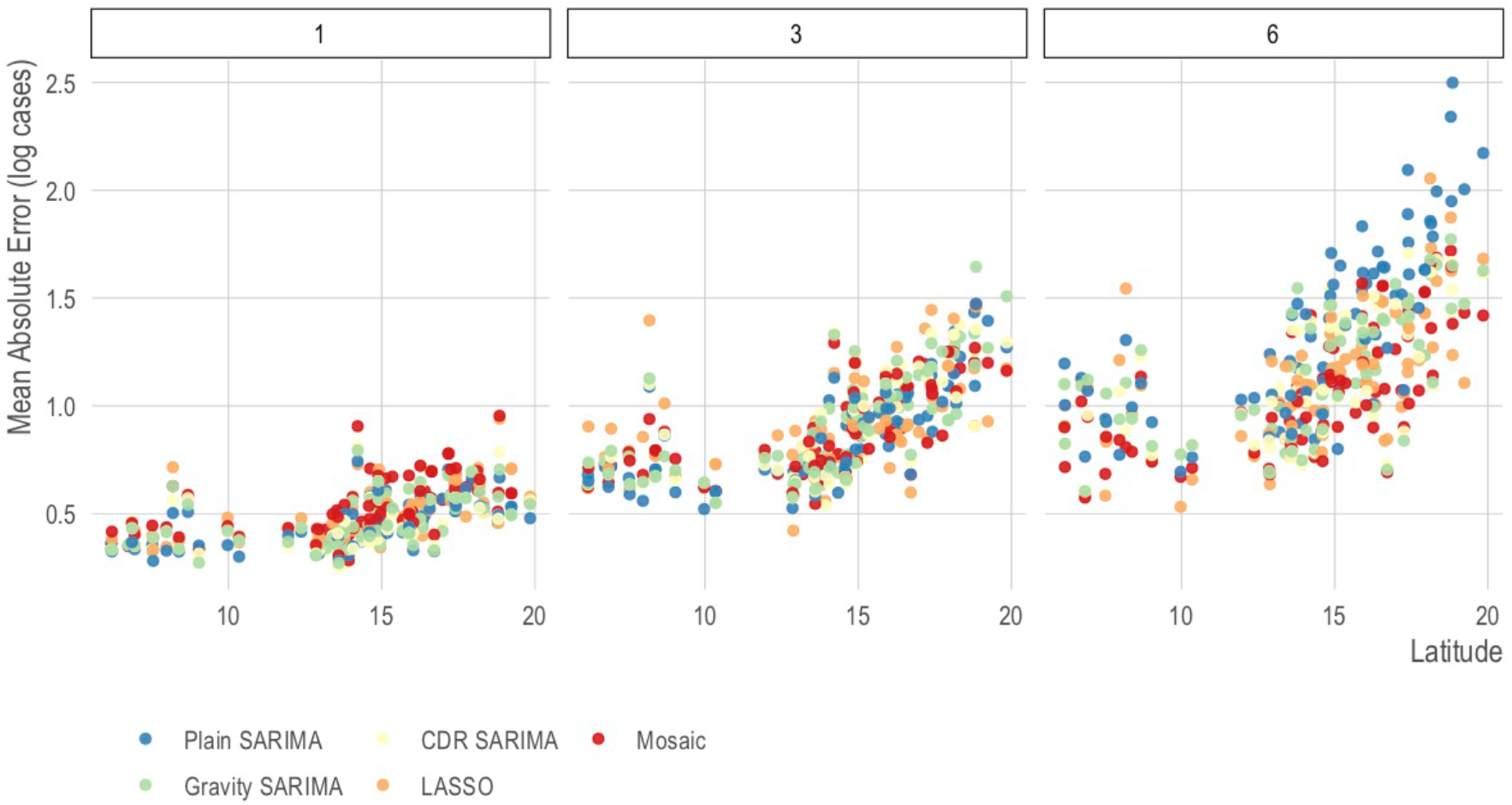
Mean absolute error for the best model in each class at t+1, t+3, and t+6 forecasting horizons for all provinces. The mean absolute error (y-axis) on the prediction (i.e., log) scale of the best model for each class for all provinces (x-axis). Provinces are ordered by latitude (x-axis, right is more northerly). There is a general decline in predictive power at farther forecasting horizons and at more northerly provinces; however, no single model or class of model performs best across all areas and all prediction horizons.

We measured the characteristics of provinces in which different models performed better or worse and found that the Plain SARIMA models performed similarly when comparing top and bottom deciles of total number of dengue cases, median number of monthly dengue cases, median monthly rate of dengue, population density, and GPP per capita. In contrast, the LASSO and mobility-augmented SARIMA models performed better in places with higher total annual cases, higher population, and lower GPP per capita (see SI Appendix, Fig. S8–S13), suggesting systematic and generalizable differences in model performance that — with more validation and in combination with geographic variation in model performance — could be used to inform model choice.

We show the feasibility of combining different classes of models by using a simple winner-takes-all voting system approach we named an adaptive mosaic model. This ensemble model selects the best performing model for each province and forecasting horizon based on the out-of-sample prediction error of previous three months, which allows the underlying base model to change over time (Figure 7). When comparing the out-of-sample prediction errors to an AR(1) model, the mosaic model outperforms the AR(1) in 107 tasks (i.e., province and forecasting horizon), underperforms the AR(1) in 3 tasks, and is not statistically significantly different from an AR(1) in the remaining 352 tasks (SI Appendix, Fig. S7). Further exploration of location-specific and task-specific voting predictions systems is outside of the scope of this study but should be explored in future research efforts.

**Figure 7.**
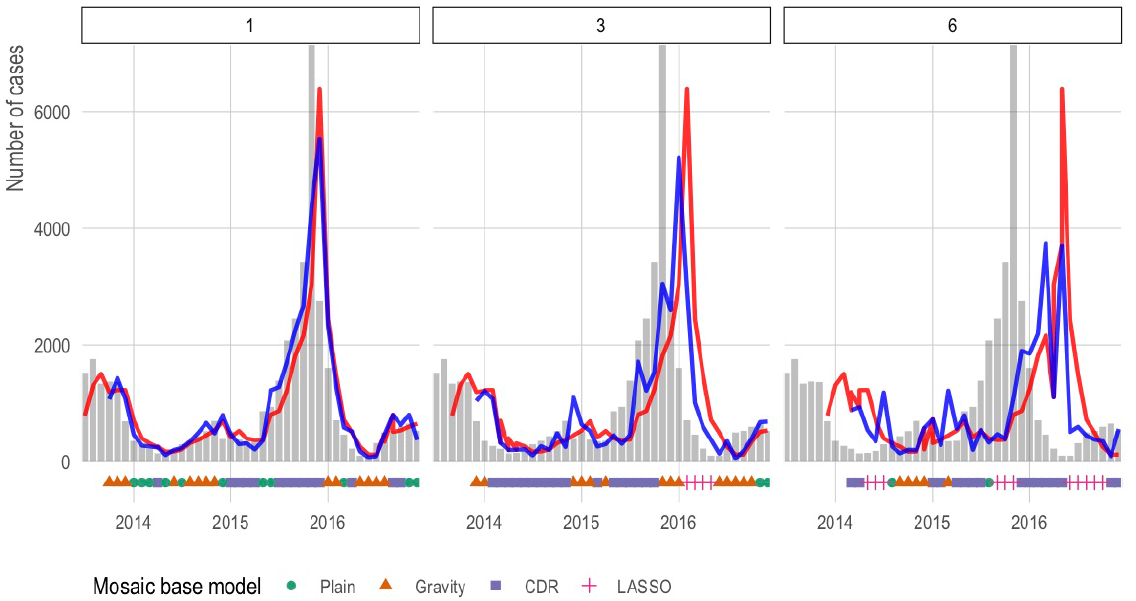
Mosaic model vs AR(1) for Bangkok at t+1, t+3, and t+6 forecasting horizons. We show the predictions for a simple mosaic model at t+1, t+3, and t+6 forecasting horizons for Bangkok in blue. For comparison, we show predictions from an AR(1) in red and observed cases in grey bars. Under each bar, we indicate the base model selected by the mosaic ensemble using a winter-takes-all approach based on the previous three out-of-sample prediction months.

## Discussion

Dengue forecasting remains an important public health challenge in Thailand and other endemic countries. Given the complexity of dengue transmission, statistical forecasting approaches like those examined here have been shown to produce meaningful disease estimates in multiple locations, and may therefore be suitable for immediate use by national control programs. In addition, we have shown that integrating additional data streams, such as information about human mobility, can improve forecasts in many areas, but the added benefit will be specific to the area and time horizon of interest. The interesting geographic variation in forecasting accuracy, which is not linked to population density or GPP per capita, may reflect the proximity to international borders with countries where frequent migration occurs. Overall, no single modeling approach can be expected to provide an optimal early warning system across all areas, even within a single country or region, or across all time horizons. So adaptive, mosaic forecasts are likely to provide the most effective approach. This type of approach could be easily integrated within the data platforms recently developed in Thailand (Nicholas G. Reich et al., 2016), which are flexible enough to accommodate different modeling approaches and forecasts.

We show that simple network methods (that implicitly incorporate human mobility) can improve upon commonly-used local SARIMA models. Also, given that the network-based approach we studied relied only on dengue case count data routinely collected by most endemic countries, we envision that similar approaches may be easily extended, and may prove to be meaningful, in many other locations around the Globe. The regularized multi-variate regression framework can also flexibly identify and incorporate additional province-level data, time lags, and other factors in the predictive model, that could be used as a hypothesis-generation tool that may capture temporal changes in inter-regional human mobility. We highlight the fact that even though the mobility data we used covered only a small fraction of time represented in the dengue case data (3.2%; i.e., 81 days vs 7 years), it was still able to improve the local (non-augmented) SARIMA, suggesting that even relatively coarse travel information would improve naïve SARIMA models. Although mobile phone data is challenging to obtain, the coarse granularity of mobility information that we used completely protects individual subscriber privacy while adding substantially to forecasting performance. Since it is continuously collected, there is no reason these data could not be aggregated by mobile operators and provided on a relatively frequent basis to disease control programs. A limitation of using CDR to model dengue transmission is that it reflects movement patterns of the entire population whereas dengue tends to occur more in children and young adults in urban areas (Limkittikul et al., 2014).

As governments prioritize how and where to spend money to improve dengue surveillance, our study suggests new regularized regression frameworks that incorporate mobility data can improve forecasts substantially. Any forecasting model will depend on the quality of the case data that it is trained upon, highlighting the primary importance of good epidemiological data. A limitation of this work is that most dengue cases in Thailand, as in most countries, are not confirmed with a diagnostic test, instead relying in syndromic surveillance. This can be unreliable with the case definition for dengue fever overlapping substantially with other causes of acute febrile illness and the completeness of the data relying on individual healthcare workers to complete the reporting forms. Thus, much of the money for better dengue forecasting should be focused on faster and better dengue case detection, more widespread diagnostic testing, sentinel surveillance of serotypes, a robust computational framework for sharing case data across regions to be analyzed centrally, and capacity building within control programs.

## Materials and Methods

### Dengue incidence data

We obtained monthly dengue case counts for over 7,000 subdistricts in Thailand from the Ministry of Public Health. These data are not available publicly and are used with the permission of the Ministry of Public Health. They consist of monthly dengue incidence counts from January 2010 through December 2016, by mutually-exclusive disease type (i.e., dengue fever, dengue shock syndrome, or dengue hemorrhagic fever). We aggregated these data to the province level and overall dengue case counts. In our data, there was a national average of 91,000 dengue cases per year with a range of 39,368 (2014) to 145,600 (2013) cases per year.

### Mobile phone data

To assess inter-province travel, we analyzed call data records (CDRs) of approximately 11 million mobile phone subscribers between August 1, 2017 and October 19, 2017. At the time of data collection, the mobile phone operator had about 26% of the market share and was the third largest provider in Thailand. In order to ensure the privacy of the mobile phone subscribers, and in compliance with national laws and the privacy policy of the Telenor group, special considerations were taken with the CDRs. First, only the mobile operator had access to the CDR and all data processing was performed on a server owned by, and only available to, the operator, thus ensuring that detailed records never left the operator or Thailand. Second, the operator provided researchers with a list of approximate cellular tower locations. For every tower location, we returned a corresponding, unlinked geographic identifier (“geocode”) of the nearest subdistrict. Mobile operator employees then aggregated the detailed CDRs up to the researcher-provided geocodes. Further spatial and temporal aggregation was performed by the researchers. These data are not publicly available and are used with the permission of Telenor Research.

To quantify travel, every subscriber was assigned a daily “home” location based on their most frequently used geocode. We tabulated daily travel between a subscriber’s home location on one day relative to the day before. Trips were aggregated to geocode-to-geocode pairs for every day and thus are memoryless — preventing the ability to trace a user (or group of users) across more than two days or more than two areas. We normalized the number of trips from geocode *i* to geocode *j* by the number of subscribers at geocode *i*. We then multiplied this proportion by the estimated population at geocode *i* to get the flow from *i* to *j*. This assumes that subscribers are more or less uniformly distributed across provinces (weighted by the population in each province). While this assumption cannot be fully tested, there is a strong correlation (Pearson’s r = .90) between subscribers and population for each province.

On average, 11.4 million subscribers (16.7% of the total population) recorded at least one event (i.e., phone call, text message, internet activity) per day (SI Appendix, Fig. S1). At both the national and provincial levels, no significant deviations from the number of subscribers or the numbers of trips occurred during this time period. For example, at the national level, the coefficient of variation for daily number of subscribers was 1.3%. Therefore, we used the mean number of trips over this time period as our estimate of inter-province travel.

### Population, gross provincial product per capita, and distance estimates

To estimate province-level population, we used the United Nations-adjusted 2015 population estimates from WorldPop (Gaughan et al., 2013), which combines remote-sensing data with other data sources to create random-forest-generated population maps. Each file contains the estimated population per pixel and was overlaid with the official administrative shapefile. We then summed the value of all pixels within each province. We used publicly available 2015 gross provincial product per capita provided by the Office of the National Economic and Social Development Board of Thailand (NESDB, 2017). The concept of “distance” is flexible in the gravity model and geodesic distance often ignores important geographical (e.g., mountain ranges) or social and behavioral constants to human mobility. In addition to calculating geodesic distance between provinces, we calculated road distance and travel time based on OpenStreetMap data using Open Street Routing Machine (Luxen and Vetter, 2011).

### Comparing observed and predicted travel

We compared observed travel between provinces with CDRs to those estimated by a gravity model with three different measures of distance: geodesic distance, road distance, and travel time. The gravity model is a popular econometric model (Tinbergen, 1963), often used to estimate mobility between areas (Lewer and Berg, 2008). The basic gravity model is:

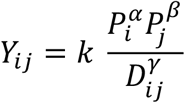

where *Y*_*ij*_ be the number of people who move from area *i* to area *j, k* is a constant term, *P*_*j*_ is the population in area *i, P*_*j*_ is the population in area *j*, and *D*_*ij*_ is some measure of distance between *i* and *j*, noting that distance may not be symmetric. The parameters *k, α, β*, and *γ* are estimated by fitting a Poisson model:

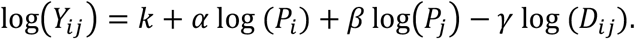

In addition to the naïve gravity model, we also adjusted for gross provincial product per capita. The best fit according to in-sample error metrics was the adjusted travel time model (SI Appendix, Table S1). We identified outlier observations as those observations with a Cook’s distance greater than five times the mean Cook’s distance.

### Quantitative methods

We evaluated the predictive accuracy of two different types of models: (1) one data-driven network approach built using an *L*_*1*_-regularized regression approach (the least absolute shrinkage and selection operator, LASSO) and (2) autoregressive integrated moving average (ARIMA) models both with and without a seasonal component (SARIMA). In addition, for the mobility-augmented autoregressive models, human mobility is accounted for by also including lagged case data from the top five areas (i.e., origins) of travelers as covariates in the model. We compared both sets of autoregressive models to the network approach predictions using a sliding window of observation and rolling forecast target as described below.

#### Network models

Based on a previous model designed to leverage spatially-correlated cases of influenza (Lu et al., 2019), we fit a multivariate linear regression on the log of dengue case counts for area *i* in month *t* with the log of dengue case counts in areas *j* at time *t* − *h* where *h* is our forecasting horizon as the covariates. Let *y*_*i,j*_ = In (*c*_*i,j*_ + 1) where *c* is the count of cases of location *i* at time *t*:

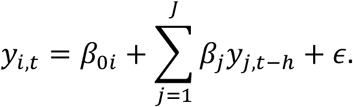

We used a sliding window of 42 months and *h* between 1 and 6. All values of *y*_*i,j*_ were standardized to be mean-centered with unit variance in order to ensure the coefficients are not scale-dependent. For all prediction months, there were more areas, 77, or input variables, than observations, 42, and thus this formulation cannot be solved using an ordinary least squares (OLS) approach. To address this, we used an *L*_1_ regularization approach to identify a parsimonious model that uses fewer variables as input than the number of available observations. This penalization approach acts to both prevent overfitting as well as selecting the most informative covariates (i.e., provinces). Specifically, we used the least absolute shrinkage and selection operator, LASSO, which minimizes the same objective function as a regular OLS while penalizing the number of non-zero coefficients with a hyper-parameter λ:

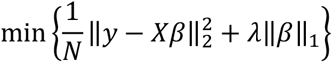

where the magnitude of the hyper-parameter λ is identified using cross validation on the training set. This approach shrinks the coefficients of non-informative or redundant areas to zero and provides for straightforward interpretation of the results allowing for identification of which areas contributed the most predictive power for any given window of observation and target area.

#### Autoregressive models

As a baseline for comparing model predictions, we used autoregressive integrated moving average (ARIMA) models, which is a common time series method applied to epidemiological modeling and dengue forecasting. These models have been used extensively in dengue prediction efforts and often incorporate a seasonal component called Seasonal ARIMA or SARIMA. Using the (*p, d, q*)(*P, D, Q*)_*s*_ convention where *p* indicates the autoregressive order, *d* indicates the amount of differencing, and *q* indicates the order of the moving average. The seasonal component, (*P, Q, D*)_*s*_, represent the same parameters with a seasonal period of *s* months. Additional exogenous variables (i.e., timeseries) can be added as covariates in this framework.

We reduced the parameter space of the SARIMA models using previous literature (Johansson et al., 2016) and our expert opinion. Specifically, we systematically search models with lags of up to four months (p = 1, 2, 3, or 4) or three years (P = 1, 2, or 3) and include a differencing order up to 1 (d and D = 0 or 1) and exclude all moving averages (q and Q fixed at 0). This results in a set of 15 model parameterizations: eight non-seasonal ARIMAs and seven seasonal ARIMAs. For each parameterization, we perform a univariate SARIMA as well as a mobility-augmented SARIMA. The mobility-augmented SARIMA incorporates the timeseries of cases from the top five connected areas, based on observed mobility, as exogenous covariates. Similar to the LASSO, we used a sliding window of 42 months, and in the case of augmented SARIMA models, we lagged the exogenous covariates by *h*.

#### Adaptive mosaic model

We show the feasibility of combining different classes of the above models by using an ensemble approach we call the “adaptive mosaic model.” For each province and forecasting horizon, we select the best performing model using a winner-takes-all approach based on the out-of-sample prediction error of the previous three months. By repeating this procedure for every prediction month, forecasting horizon, and province, the underlying base model can adapt over time (Figure 7).

#### Accuracy metrics and model comparison

Consistent with previous research (Lauer et al., 2018; Nicholas G Reich et al., 2016), when assessing predictive performance of a single model, we used mean absolute error (MAE) and when assessing the relative performance of two models, we used relative mean absolute error (relMAE). The MAE of the log transformed counts is as follows:

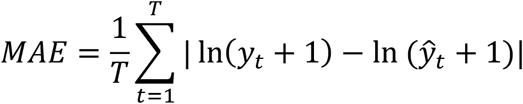

where *y*_*t*_ and 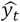 are the observed and average counts for prediction month *t*. One strength of this approach is that the MAE will be the same regardless of magnitude as long as the ratios are the same (i.e., 100 and 110 for predicted and observed will result in 1.1, just as 10 and 11 or 11 and 10). This is an important feature given the differences in population size and case counts between provinces.

When comparing model *A* to model *B* at forecast horizon *h*, we take the ratio of their MAEs:

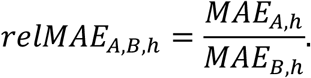

To assess the predictive performance of each model, we used retrospective out-of-sample estimates of the mean absolute error, assuming we only had data prior to the time of estimation and based on a 42-month sliding window of observation. For example, the 6-month prediction for June for one year would only include data up to December for the year before and only as far back as 42 months from that December. This provides 42 months of evaluation data or up to 41 separate models to evaluate prediction error (noting that the number of months available in the evaluation period is also a function of the prediction horizon). To compare across multiple models (e.g., to find the model with the best t+1 month forecast in a single province), we used the baseline AR(1) (i.e., ARIMA(1,0,0)(0,0,0) with no exogenous variables) as our referent model. Thus, the relMAE can be interpreted as the relative under- or over-performance of our model compared to a standard epidemiological model, averaged over all prediction months.

To assess the utility of call detail records, for each province and forecasting horizon we selected the best performing model of each class. We then compared the CDR SARIMA to each other class using a Wilcoxon signed-rank test to compare the out-of-sample prediction errors. Statistically significant differences are shown in the province-specific reports (SI Appendix, Text S1) and in Figure S7. Similarly, we compared the proposed mosaic model to a simple AR(1) using a Wilcoxon signed-rank test (SI Appendix, Fig. S8).

## Data Availability

Raw case data are provided by the Ministry of Health and cannot be shared without a data use agreement. Raw mobility data are provided by Telenor Research and cannot be shared without a data use agreement.

## Acknowledgements

RJM and NE were supported by Asian Development Bank TA-8656. The content is solely the responsibility of the authors and does not necessarily represent the official views of the funders. MS was partially supported by the National Institute of General Medical Sciences of the National Institutes of Health under Award Number R01GM130668. The content is solely the responsibility of the authors and does not necessarily represent the official views of the National Institutes of Health. CB and MS thank the Harvard Data Science Initiative for their support in partially funding this collaborative work.

## Author Contributions

CB conceptualized the study. MVK, CB, and MS designed the methodology. MVK conducted all analyses. MVK prepared the original draft. All authors provided critical feedback. KE-M, NE, DA, and RJM curated the data. MVK, MS, JTC, NK, and CB interpreted the results. CB and RJM supervised this work. All authors reviewed and approved the submitted manuscript.

